# Trauma-informed approaches in primary healthcare and community mental healthcare: a mixed methods systematic review of organisational change interventions

**DOI:** 10.1101/2022.07.09.22277443

**Authors:** Natalia V Lewis, Angel Bierce, Gene S Feder, John Macleod, Katrina M Turner, Stan Zammit, Shoba Dawson

## Abstract

A trauma-informed approach is a framework for organisational (synonym system) change interventions that address the universal prevalence and impact of trauma. This mixed methods systematic review assessed the effects of trauma-informed approaches on psychological, behavioural, and health outcomes in healthcare providers and adult patients in primary care and community mental healthcare. We searched five databases and grey literature and consulted experts for reports published in January 1990-June 2021. The quantitative descriptive and qualitative framework syntheses were integrated through a line of argument and mapped onto a logic model. We included six non-randomized studies that evaluated eight interventions with varied theoretical development, components, and outcomes. The most common components were budget allocation, workforce development, identification/response to violence and trauma, and evaluation. Evidence for intervention effects was limited and conflicting. Four studies reported improvement in provider readiness and sense of community, while three reported conflicting effects on provider behaviour regarding delivery of trauma-informed care. Four studies reported some improvement in patient readiness for disease management and access to services; however, the evidence for patient satisfaction was conflicting. Two studies found that patients and providers felt safe.

While one study reported improvement in patient quality of life and chronic pain, another found no effect on substance use, and three studies reported conflicting effects on mental health. Interventions mechanisms included a package of varied components, tailoring to the organisational needs, capacities, and preferences, staff education and self-care, creating safe environments, shared decision-making. Intervention effects were moderated by contextual (health system values, policies, governance, business models, trauma-informed movement, organisational culture, social determinants of health) and intervention factors (buy-in from all staff, collective learning through conversations, equal attention to staff and patient well-being, sustainable funding). No studies measured adverse events/harm, cost effectiveness, or providers’ health. We need more methodologically robust evaluations of trauma-informed organisational change interventions.

A preprint of this article has previously been deposited in the preprint server for health sciences [1].

## Introduction

Psychological trauma has devastating impact on the health of individuals, communities, and societies [2]. Traumatic experiences can be caused by single events (e.g., sexual assault, unexpected family death) or chronic phenomena (e.g., adverse childhood experiences [ACEs], domestic abuse, community violence, historical trauma) [3]. Structural inequalities (e.g., healthcare, economic, gender, and racial disparities) may exacerbate effects of these traumatic experiences [4].

Lifetime traumatic events are associated with risk-taking behaviours, poor health, adverse socio-economic outcomes, and increased use of primary care and mental health services [2]. Coercive practices and invasive procedures within healthcare services (e.g., removal of choice regarding treatment, judgemental attitudes following a disclosure of abuse, lack of accessible services) can re-trigger or re-traumatise both patients and healthcare staff [5]. As a result of empathetic engagement with trauma survivors, healthcare providers can experience secondary traumatisation and/or vicarious trauma [6].

Over the last two decades, a trauma-informed approach has gained momentum as a framework for organisational (synonym system) change interventions that address the high prevalence and impact of trauma among healthcare providers and users. The approach differs from standard ‘trauma blind’ service delivery by integrating 4 *Rs* throughout healthcare organisation: **r**ealising and **r**ecognising the impacts of trauma on patients and staff, **r**esponding by integrating knowledge about trauma into policies and practices, and creating environments and relationships that prevent **r**e-traumatisation and promote physical and emotional safety for all [7]. The framework of a trauma-informed approach is not a protocol but high-level guidance for organisational change interventions that can be adapted to any health service. Although different authors used differing terminology and definitions, they mostly aligned with the philosophy and principles of the trauma-informed approach proposed by Harris and Fallot [8] and developed further by the US Substance Abuse and Mental Health Services Administration (SAMHSA) [9]. Subsequent framework developments drew attention to the intersection of individual and interpersonal trauma and structural inequalities [5, 10, 11], universal applicability of the trauma-informed approach [12], benefits to patients and staff [13], and application to services other than mental health and addiction [11, 14] (Supplementary material Table S1).

Its proponents consistently highlighted the organisational level of a trauma-informed approach, requiring changes in the structure and culture of the organisation (organisational domain). These organisational changes should precede changes in clinical practices (clinical domain) [8]. Becoming a trauma-informed organisation is described as a transformation process rather than a one-off activity. The transformation work is guided by the SAMHSA six key principles of safety, trust, collaboration, choice, empowerment, and cultural sensitivity. These principles can be implemented through varied intervention components and activities tailored to organisational needs, abilities, and preferences and to the wider contexts [9]. One contested component is screening for a history of traumatic events in adult healthcare settings [15]. Most authors consider it an essential component [8-10], while some think that disclosure of violence and trauma is not the goal of a trauma-informed approach and service providers do not necessarily need to know about peoples’ lived experiences to provide appropriate healthcare [13]. The conceptual mutability of a trauma-informed approach framework and lack of empirical evidence for effectiveness has been challenged [16]. These and the various definitions and applications might have contributed towards misconceptions about trauma-informed approaches at the organisational level, for example, confusion between universal trauma-informed organisational change interventions for all staff and patients and trauma-specific treatments for people with consequences of trauma [17].

A growing body of literature, policies and guidelines recommend trauma-informed approaches in healthcare organisations and health systems; however, the evidence base for the effectiveness is still being assessed [18-21]. Our pilot searches and consultations with experts found extensive literature on articulating trauma-informed approaches, and how and why we should embrace and evaluate them. We identified a growing market of training and certification on trauma-informed approaches. In contrast, we found a small number of evaluations of the effectiveness of trauma-informed organisational change interventions within healthcare. Currently, studies of standalone training interventions about trauma-informed care without any changes at the organisational or wider system level dominate the evaluation literature [22]. While a few evaluations of trauma-informed organisational change interventions were conducted in secondary mental healthcare [23] and services for children [24, 25], we found no systematic reviews of the trauma-informed approach in adult primary care and community mental healthcare. These services are a patient’s first point of contact with a health system [26].

This systematic review is part of a programme of research on trauma-informed health systems (TAP CARE study). We aimed to systematically identify, appraise, and synthesise the empirical evidence on trauma-informed organisational change interventions in primary healthcare and community mental healthcare to understand:

1. What models of trauma-informed organisational change interventions have been applied?
2. What are the effects of these interventions on psychological, behavioural, and health outcomes in healthcare providers and adult patients?
3. Are these interventions cost-effective?
4. What programme theories were proposed to explain intervention effects?

## Materials and methods

### Design

We registered study protocol with PROSPERO (CRD42020164752) and have published it elsewhere [27]. In brief, we conducted a mixed methods systematic review with a results-based convergent synthesis [28]. The authors’ positionality within the critical realism paradigm [29] influenced decision to treat quantitative and qualitative findings equally and do not undertake data transformation. This report follows the PRISMA 2020 statement [30, 31].

### Patient and Public Involvement

In line with the key principles of a trauma-informed approach [9], we involved people with lived experience of trauma in each stage of the review. The public advisory group included eight people with lived experience of trauma. The professional advisory group included ten people who plan, fund, and deliver health services. Both groups discussed research questions and listed outcomes that they viewed as meaningful to patients, service providers, managers, and funders of services. The professional group also developed a list of UK primary and community mental health services. We met with the advisory groups every six months to consult on data extraction, logic model refinement, interpretation, and dissemination of study findings.

### Development of a logic model

We used the measurement model for trauma-informed primary care [32] as a foundation for our logic model and incorporated data from background literature in version 1 [27]. Version 2 incorporated findings at the data extraction stage. Version 3 incorporated revisions at the data synthesis stage and included the following constructs:

- A component of the trauma-informed organisational change intervention categorised by the ten SAMHSA implementation domains [9].
- An intermediate psychological (cognitive or affective) or behavioural outcome regarding trauma-informed care that the components might influence (e.g., provider readiness or practices) categorised by the four-level framework of the healthcare system (individual patient, care team, organisation, political and economic environment) [33].
- A long-term health-related outcome/phenomenon of interest that the intermediate outcomes/phenomenon of interest might influence (e.g., patient or provider mental health) categorised by the four-level framework of the healthcare system [33].
- A moderator – a factor that could affect either positively or negatively, the link between a component and any outcome (Figure 1).

**Figure 1.**
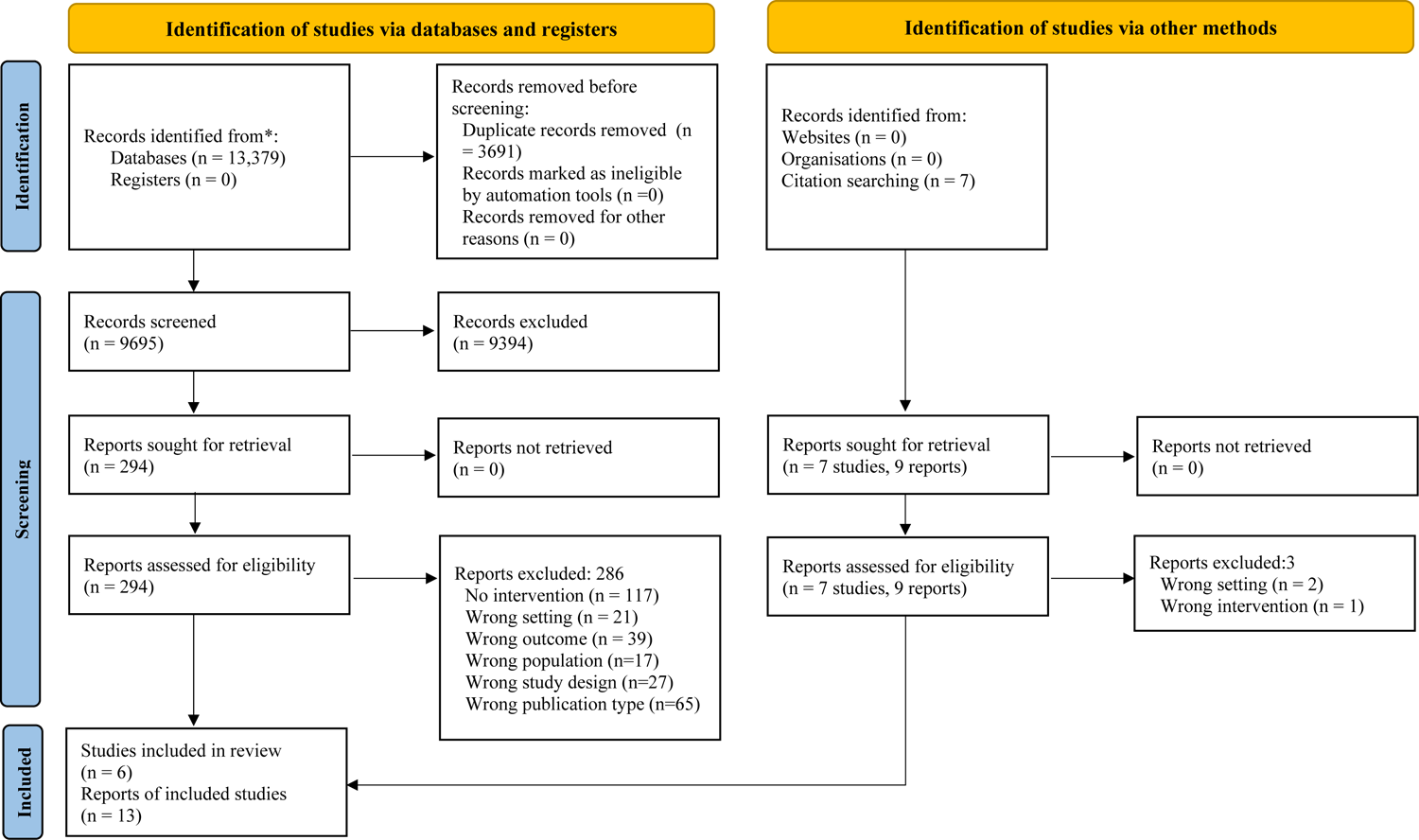
PRISMA 2020 flow

### Search strategy and selection criteria

Based on previous systematic reviews [34, 35] and the expertise of the research team and advisory groups, the first reviewer (SD) developed a search strategy combining MeSH and free-text terms. SD conducted scoping exercises in different databases to maximise the search strategy’s sensitivity and specificity. The search terms were modified and tailored for five electronic bibliographic databases: Cochrane Library, MEDLINE, EMBASE, Cumulative Index to Nursing and Allied Health Literature (CINAHL) and PsycINFO. We limited the search to primary studies published between January 1990 and February 2020, updated in June 2021 (Supplementary material Table S2).

SD searched the PROSPERO database for relevant systematic reviews in progress, and the ethos library and PROQUEST for dissertations. Additionally, SD conducted a grey literature search on websites of organisations involved in development and application of trauma-informed approaches. SD and NVL checked references and citations of included papers. NVL contacted corresponding authors, subject experts in trauma-informed approach, and study advisory groups for additional reports. We included primary studies of any design that evaluated a trauma-informed organisational change intervention in primary care or community mental healthcare (Table 1).

**Table 1.**
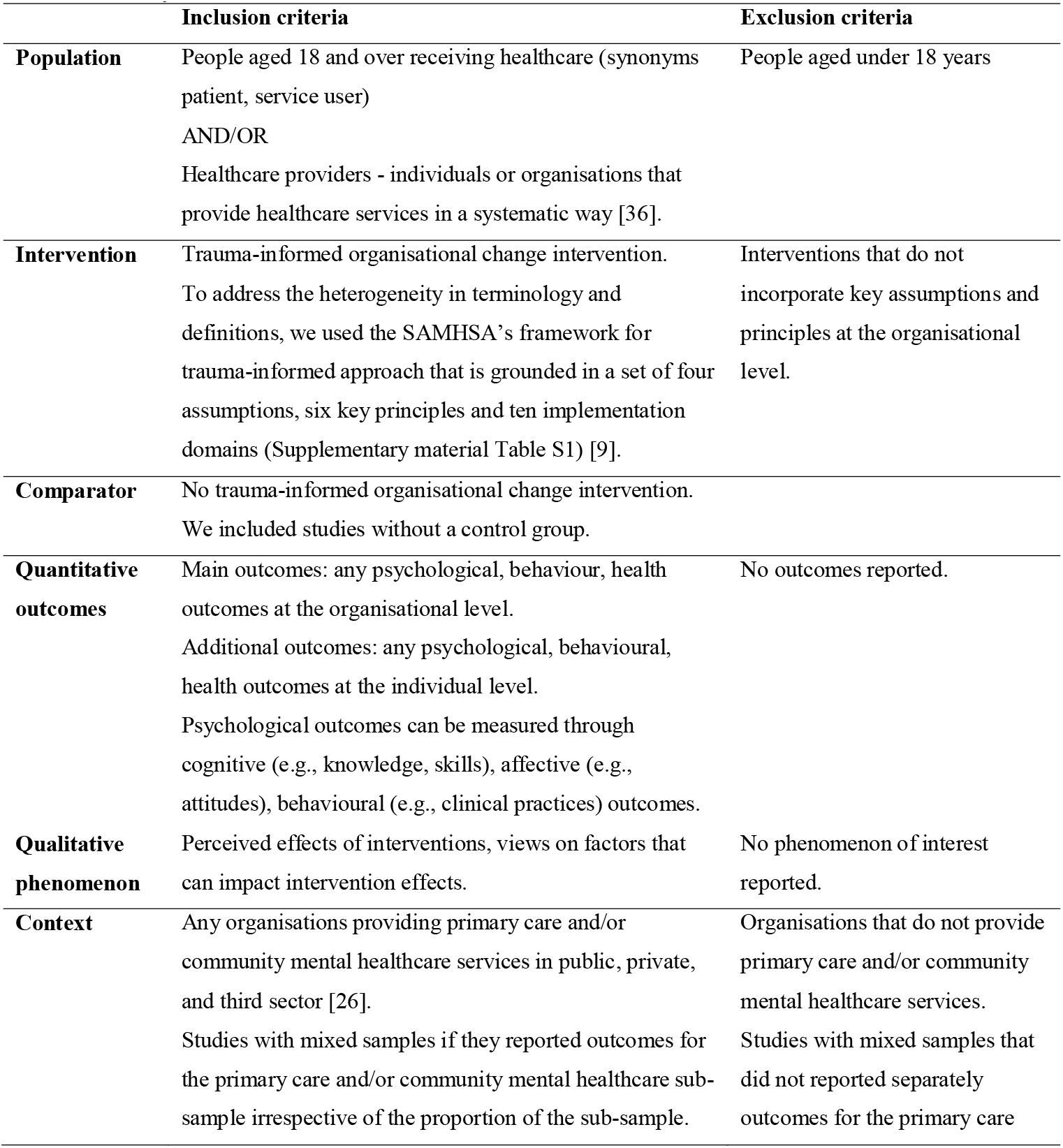
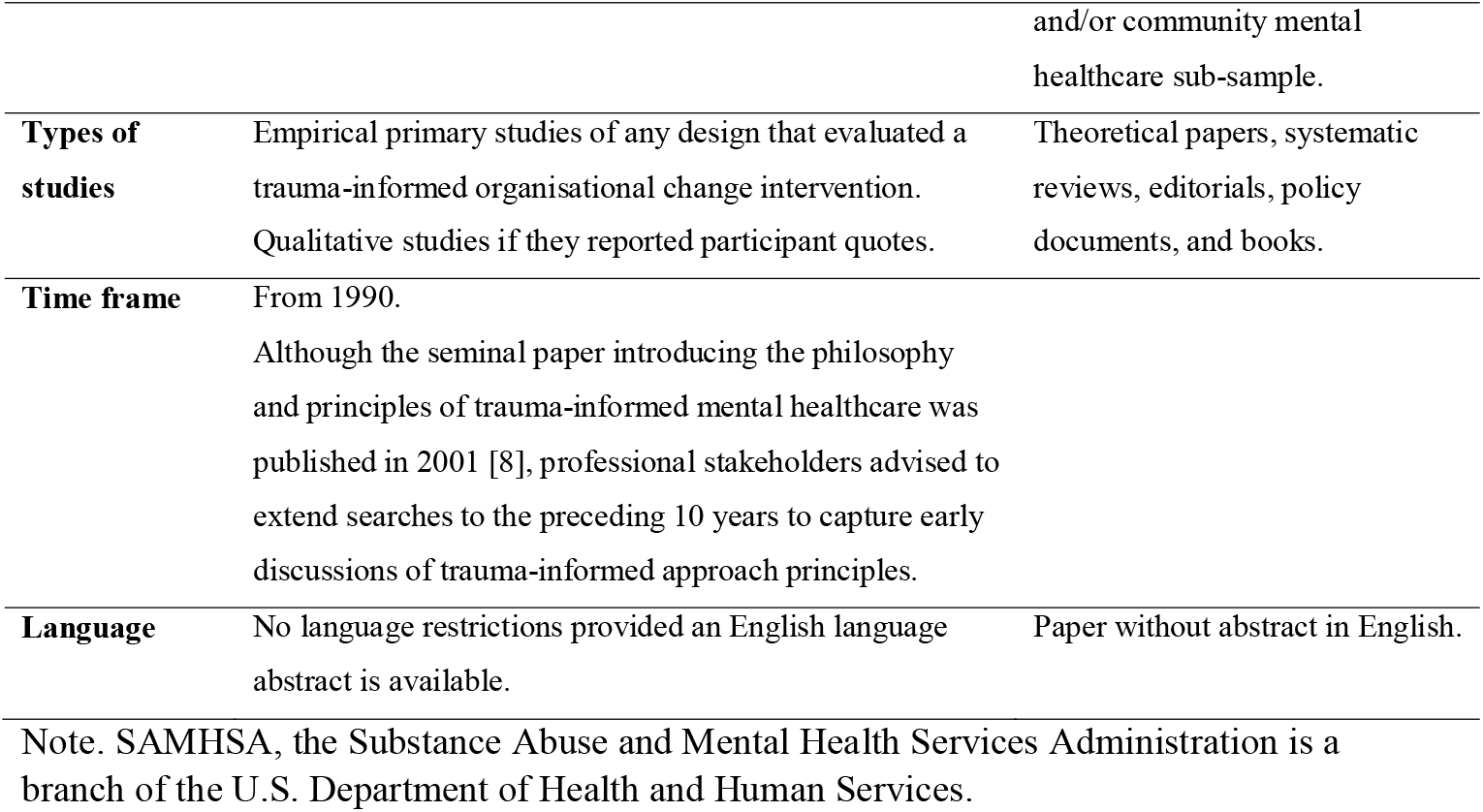
Study inclusion and exclusion criteria

### Study selection

We used Rayyan [37] to combine, export, and screen the results of the database searches. The first reviewer (SD) and a second reviewer (AB or NVL) independently screened titles and abstracts and full reports against study inclusion criteria (Table 1). The reviewers met and resolved discrepancies through discussion. Where they could not reach consensus, senior team members (GF, JM) acted as third reviewers. We included multiple reports of the same study if they contained new information and collated multiple reports so that each study was the unit of analysis. The earliest most detailed report was used as study ID.

### Data extraction

SD adapted a data extraction proforma from previous systematic reviews. For each quantitative outcome, we extracted type of measure and effect estimates as reported in the primary study. If a follow-up measure was reported repeatedly, we extracted all results. If a study recruited a mixed sample or had multiple sites, we only extracted data relevant to adult primary care or community mental healthcare. SD extracted data and NVL checked and reconciled the forms and asked all corresponding authors to check. Five of the six authors responded.

We treated included reports as primary qualitative data and used NVivo 12 to simultaneously extract and code data on intervention characteristics and qualitative phenomena of interest (i.e., perceptions of intervention effects or factors that might influence intervention effects). We used the framework synthesis method recommended for addressing applied policy questions [38]. Our initial coding frame included constructs from the SAMHSA framework of trauma-informed approach [9], our logic model, and the four-level healthcare system model [33]. Two reviewers (NVL, KT) deductively coded intervention description, participants’ quotes, and authors’ interpretations relevant to our research questions. First, NVL and KT independently manually coded two reports and met to discuss the codes. Then NVL imported the framework into NVivo and completed the coding, refining the framework throughout this process and grouping codes into themes.

### Quality appraisal

We conducted quality appraisal as part of data extraction to indicate methodological limitations in each included study. Since we included studies of multiple designs, we used the Mixed Methods Appraisal Tool (MMAT) [39]. SD completed the MMAT checklist for each study, NVL checked and reconciled through consensus.

### Data synthesis

We conducted a results-based convergent synthesis [28] at three stages: (i) concurrent quantitative and qualitative syntheses, (ii) integration of findings from the two syntheses through a line of argument and (iii) mapping onto a logic model [40]. At stage one, SD synthesised quantitative results in tables and descriptive summaries. Due to the variation of intervention models, measures, and outcomes, we could not conduct a meta-analysis. NVL grouped deductive codes into themes and wrote descriptive accounts with illustrative quotes. At stage two, NVL and SD displayed quantitative and qualitative syntheses in tables and developed lines of argument for integrated outcome domains, intervention mechanisms, and moderators. We judged intervention effects by change in any quantitative outcome and/or participant perception of change reported in the primary studies. In the quantitative synthesis, we used authors’ interpretation of their results based on p values, 95% confidence intervals (CI), or point estimates. In the qualitative synthesis, we summarised participants’ quotes and authors’ interpretations of primary data about perceived intervention effects. We categorised measured and perceived effects as *improvement, mixed effect, nil effect, negative effect/harm*. We ascribed a *mixed effect* when one or more, but not all measures of the same outcome changed under the same intervention. If different studies reported contradicting findings on the same outcome, we categorised such evidence as *conflicting*.

At stage three, NVL mapped the integrated lines of argument onto the constructs of the logic model. The final logic model only included items that were supported by evidence from the included studies (Figure 1).

## Results

We included 13 reports [41-53] of six studies [41, 42, 45, 48, 50, 52] (Figure 2).

**Figure 2.**
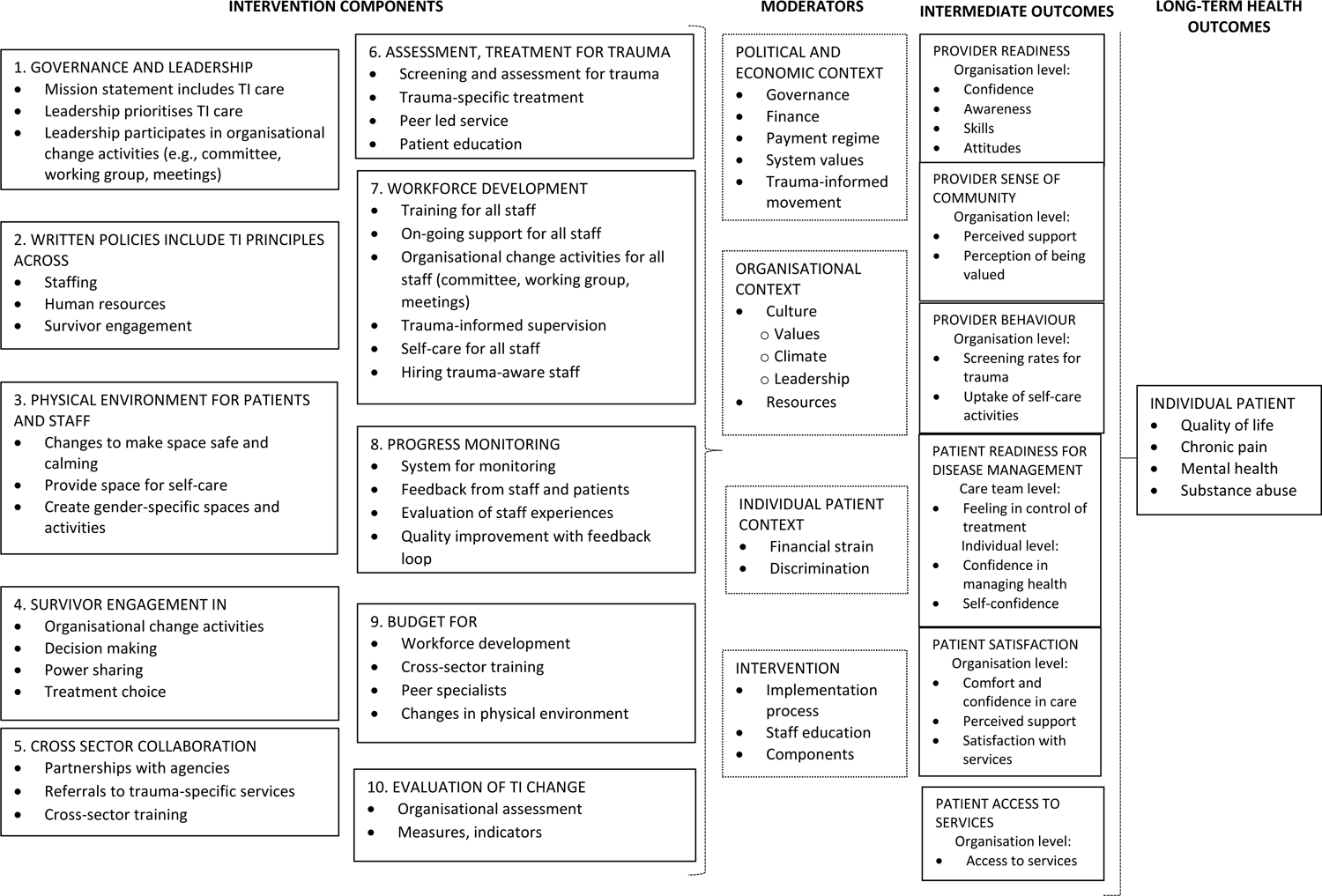
Logic model

### Study characteristics

The primary studies used non-randomised quantitative and qualitative designs. Of six studies, three were from the US [48, 50, 52] and one each from the UK [41], Canada [45] and Australia [42]. Three studies were service evaluations [41, 42, 48], two assessed new organisational change interventions [45, 52], and one was a quality improvement project [50]. The quantitative studies/components used controlled [52] and uncontrolled [45] before-after design and cross-sectional design [50]. Only two studies reported follow-up measures at 12 [52], 18 and 24 months [45]. Qualitative studies/components used focus groups, interviews, and observation [41, 42, 48].

Trauma-informed organisational change interventions were applied in public primary care clinics that served populations with high rates of trauma [45, 48, 50] and public [42, 52] and third sector [41] specialist organisations that served women with a history of interpersonal violence. Two studies were single-site evaluations [41, 42] and four were multi-site studies [45, 48, 50, 52]. The Equipping Primary Health Care for Equity (EQUIP) study took place in four Canadian primary care clinics that used the same intervention model [45]. From the US Women Co-occurring Disorders and Violence Study (WCDVS), we included the Washington DC site with four community mental health centres [52]. From the Dubay et al report [48], we included three primary care settings that used different intervention models: Women’s HIV Clinic San Francisco, Montefiore Medical Group of 22 primary care practices New York, and Family Health Clinic Philadelphia. From the US Aspire to Realize Improved Safety and Equity (ARISE) evaluation [50] and Australian Young Women’s Clinic [42], we extracted data for the patient group aged 18 and above.

The total number of participants in the included studies was 117,447 patients and 137 healthcare providers. The number of patients ranged from 6 in qualitative service evaluation [48] to 116,871 in analysis of routine data [50]. The number of providers (nurses, physicians, counsellors, outreach workers and allied professionals) ranged from 4 [41] to 117 [45] (Table 2).

**Table 2.**
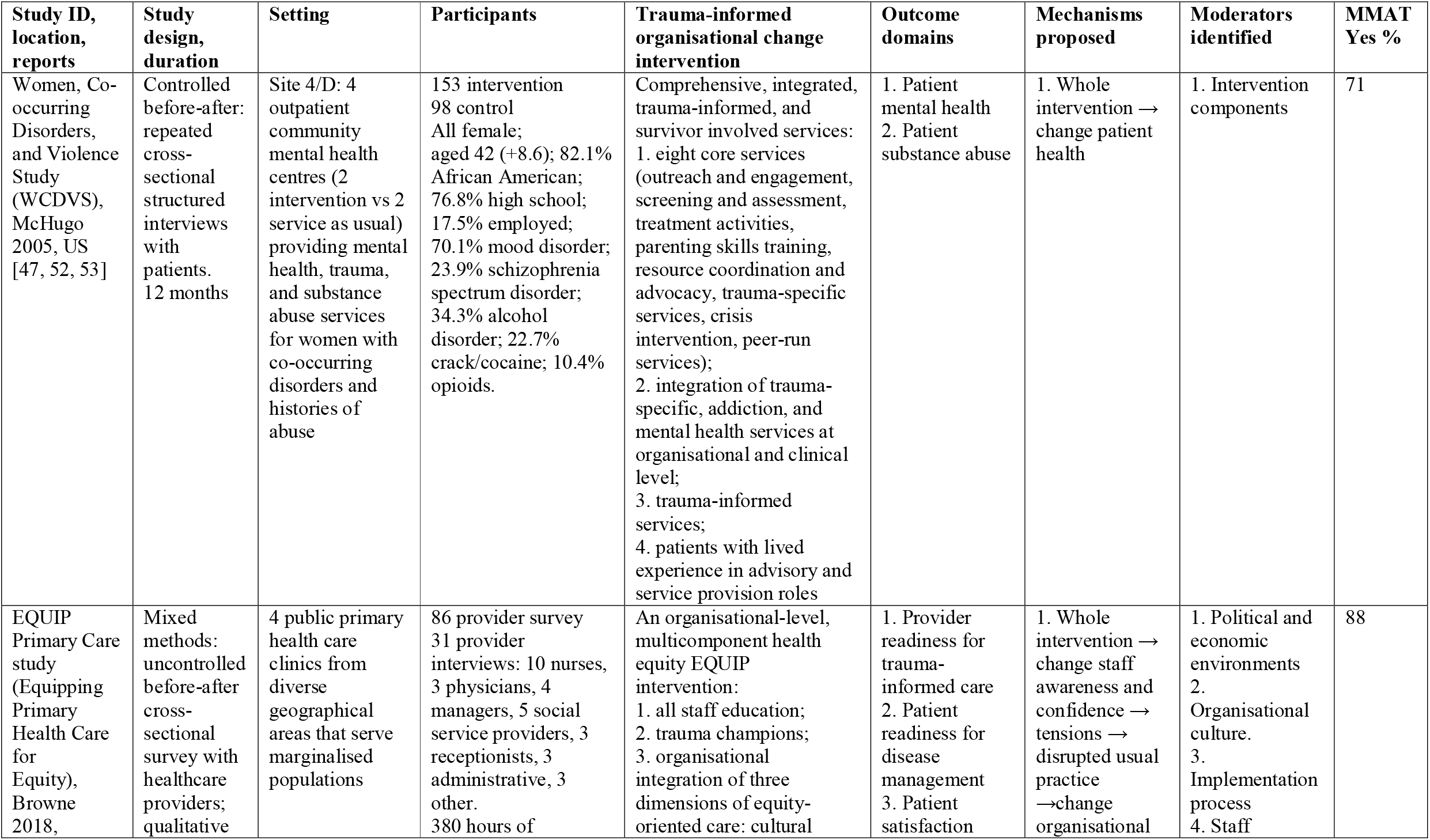
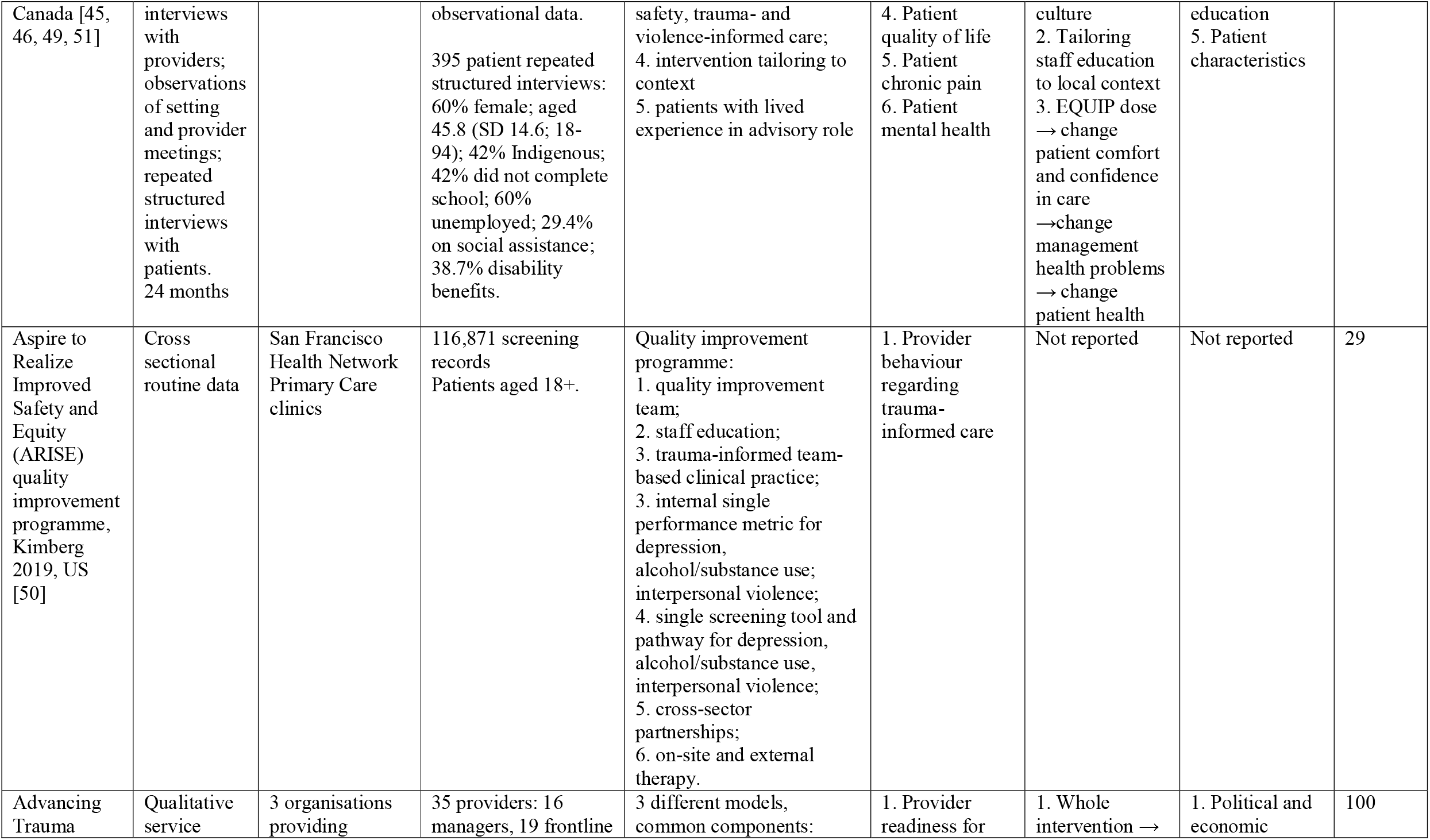
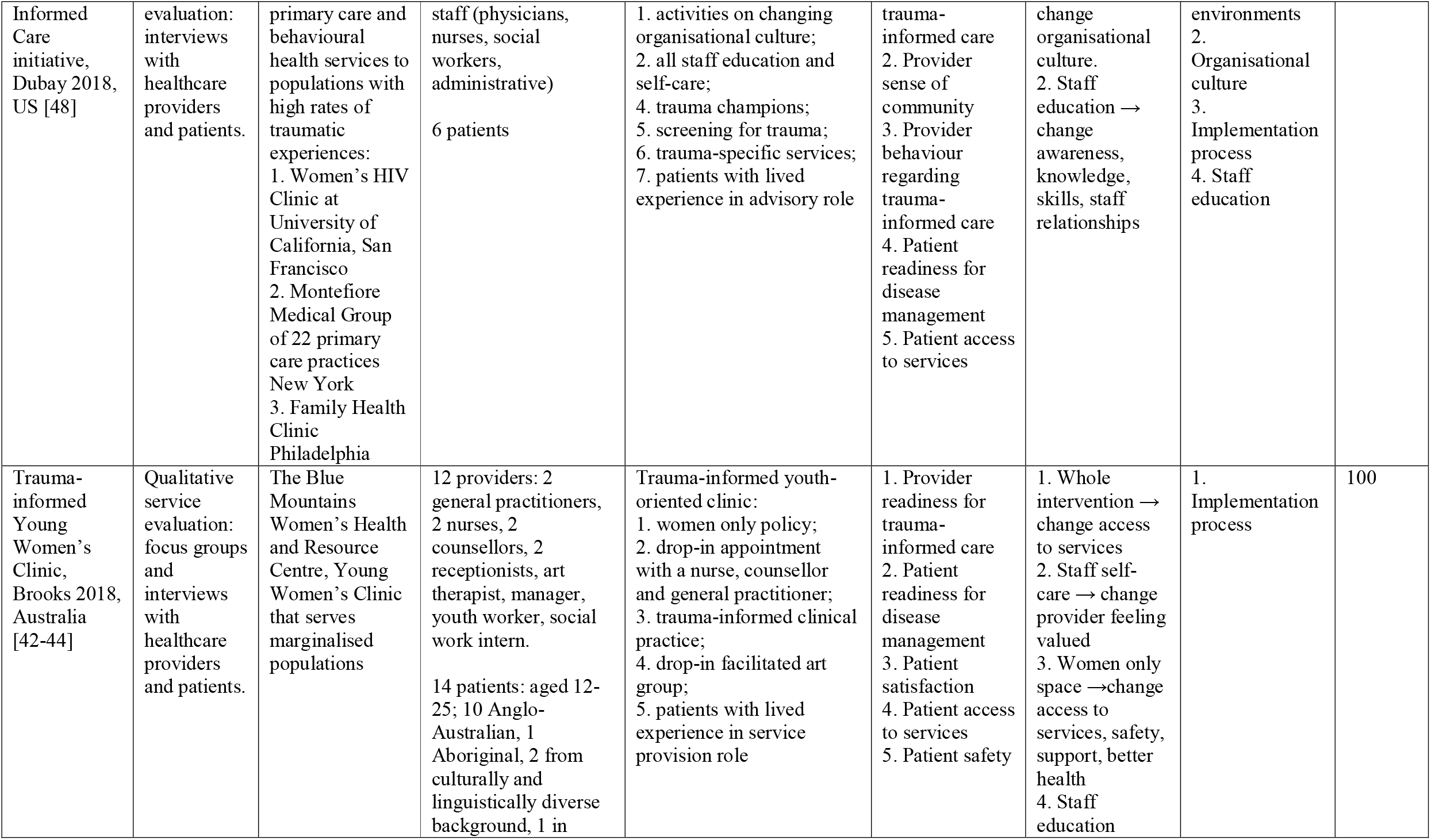
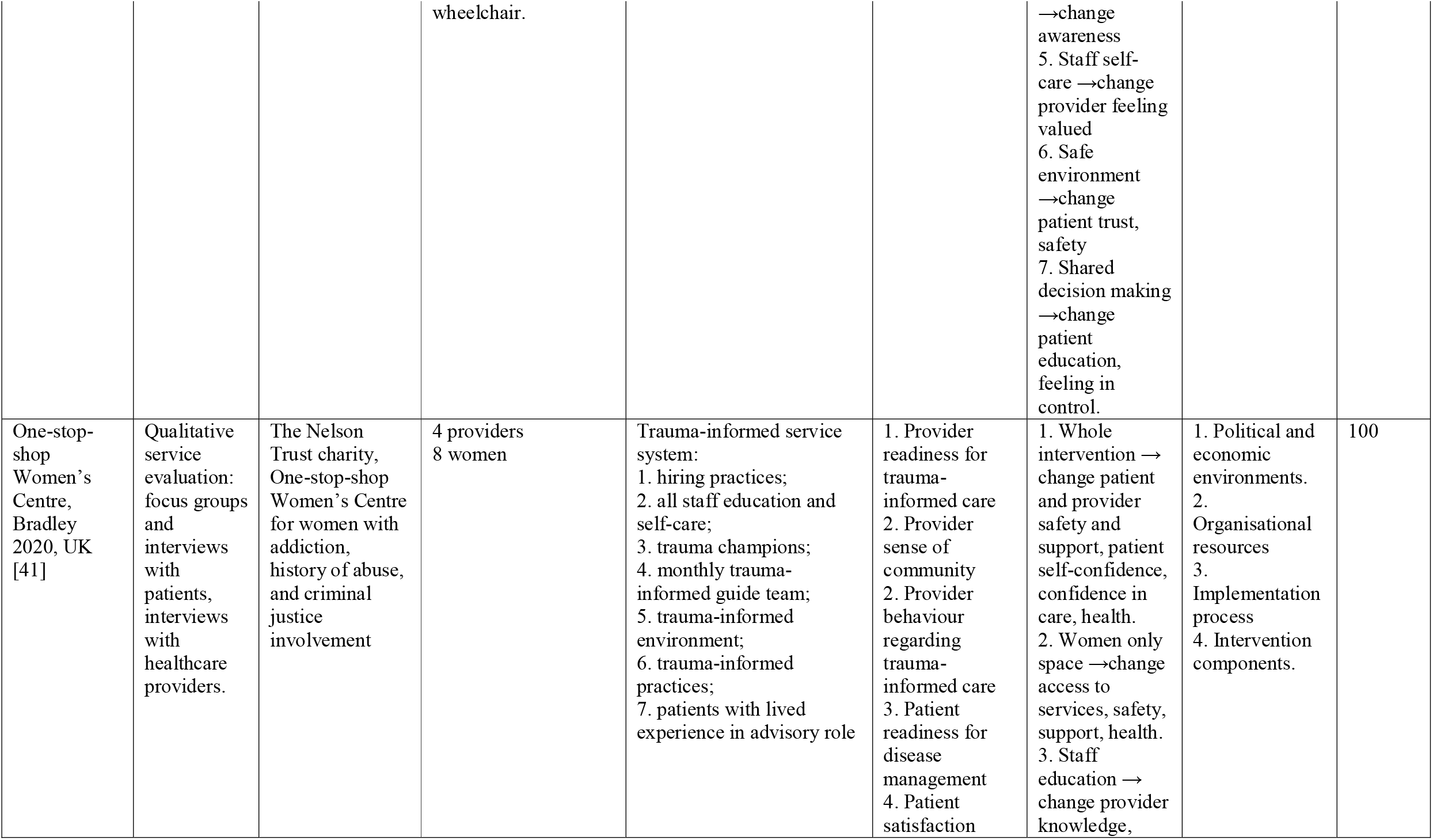
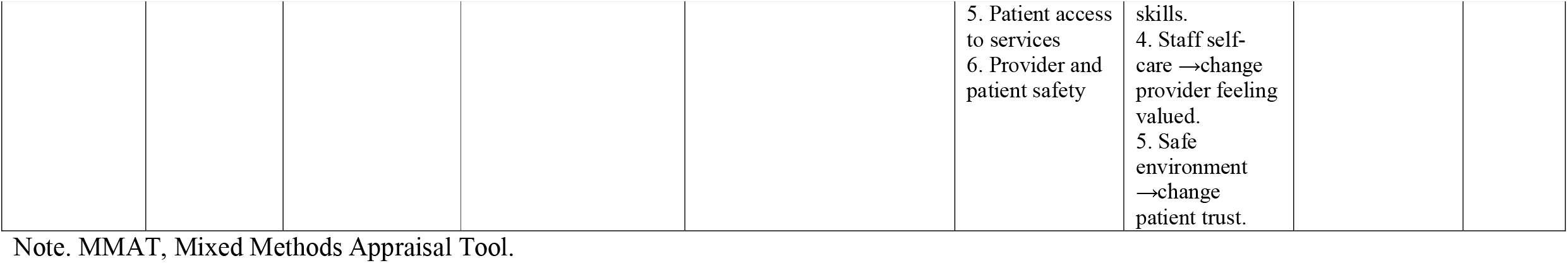
Study characteristics

### Models of trauma-informed organisational change interventions

The six studies evaluated eight different models of trauma-informed organisational change interventions. Although each model was tailored to the patient population, organisation, and wider contexts, all the models had sufficient common features for cross-study comparison. Our framework synthesis confirmed that each model aligned with the *4Rs* of the SAMHSA framework. The models varied by their level of theoretical development, formalisation, and activities within each component (Supplementary material Table S3). Three of eight interventions used existing models of trauma-informed organisational change interventions. The UK One-stop-shop Women’s Centre had been using the Trauma-informed Service Systems model [8] for less than a year [41]. The Family Health Clinic Philadelphia had been adopting the Sanctuary Model [54], while the Women’s HIV Clinic San Francisco had used the Trauma-informed Primary Care framework [55] for more than two years [48]. The Canadian team developed and evaluated the new EQUIP intervention for 24 months [45]. The other four sites (WCDVS Washington site, Montefiore Medical Group, Young Women’s Clinic, San Francisco Health Network Primary Care) applied tailored organisational change interventions up to 12 months [52], 17 months [50], more than 24 months [48] to 13 years [42]. The Sanctuary Model by Bloom has been operationalised as a certified business model [54].

The intervention components varied in the extent to which they mapped onto the SAMHSA ten implementation domains [9] (Supplementary material Table S3). Only EQUIP [45] and Trauma-informed Young Women’s Clinic [42] included components from all ten implementation domains; the Montefiore Medical Group intervention covered six domains [48]. The four common domains across all eight models were: (i) budget, (ii) workforce development, (iii) identification and/or response to violence and trauma, and (iv) evaluation of change. All interventions were funded through project grants or joint financing. The budgets covered training and ongoing support for all staff, trauma-informed practices, and changes in the physical environment. The content, format, and duration of the training varied; the common features were delivery by external experts, tailoring to the organisational context and patient population, and booster sessions. Similarly, tailored self-care activities included mindfulness sessions, wellbeing days, and trauma-informed supervision. All models included on-site, or external trauma-specific treatments tailored to the population served.

Five interventions included screening for history of trauma and/or mental health conditions [41, 48, 50, 52]. Seven models made changes in the physical environment. These ranged from women-only spaces and activities [41, 42, 52] with provision of childcare [41, 42], through redesigning waiting rooms and offices [41, 45, 48] to extending opening time [45] and consultation length [42]. Varied activities to monitor progress and quality were reported in seven models [35, 36, 39, 42, 44]. Cross-sector collaboration was reported in six models [41, 42, 45, 48, 50, 52]. Engagement and involvement of people with lived experience in organisational change were reported in six models [41, 42, 45, 48, 52]. Organisations undertook different engagement activities, from including people with lived experience of violence and trauma in working groups/committees [48] to hiring them as advisers [45, 50] and service providers [41, 52]. The least common domains were the leadership and governance support in five models [41, 42, 45, 48, 50]. Written policies and procedures that reflected commitment to a trauma-informed approach were reported in three models [42, 45, 50] (Supplementary material Table S3).

### Methodological quality of included studies

The quantitative non-randomised studies and mixed methods study components were of moderate quality. The methodological quality of qualitative study components was high (Table 2). Most studies had clearly defined research questions, which were addressed by the data collected [41, 42, 45, 48, 52]. One study did not pose a clear research question [50]. All four qualitative studies/component showed coherence between data sources, collection, analysis, and interpretation [41, 42, 45, 48]. The three quantitative studies/component used appropriate sampling techniques and measures [45, 50, 52]; two had a good completion rate [45, 52]. Only one study considered possible confounding factors and confirmed that the intervention was administered as intended [45]. The only mixed methods study provided a design rationale and adequately integrated the qualitative and quantitative components [45]. However, it did not address the divergence or inconsistency between components, nor did it fulfil the methodological quality criteria (Supplementary material Tables S4, S5).

### Effects of trauma-informed organisational change interventions on patient or healthcare provider psychological, behavioural, and health outcomes

We found limited and conflicting evidence for the effects (or perceived effects) of trauma-informed organisational change interventions on 11 outcome domains, with an overall direction towards some improvement. Most evidence came from the controlled before-after WCDVS study [52], mixed methods EQUIP study [45], and three qualitative service evaluations [35, 36, 42]. The evidence for each intervention model was based on a single study. None used the same measures (Table 3, Figure 1).

**Table 3.** Evidence for effects and perceived effects of trauma-informed organisational change interventions on patients and provider outcomes

| Outcomes/phenomena of interest domain | Nil effect |  | Improvement |  | Mixed effect |  |
| --- | --- | --- | --- | --- | --- | --- |
|  | Quantitative | Qualitative | Quantitative | Qualitative | Quantitative | Qualitative |
| **INTERMEDIATE COGNITIVE, AFFECTIVE, AND BEHAVIOURAL OUTCOMES** |  |  |  |  |  |  |
| 1. Provider readiness for trauma-informed care | None | None | **Organisation level**<br>1.1. Confidence scores 5.8 (SD 2.02) →6.9 (SD 2.00) →7.7 (SD 1.34) [45] | **Organisation level**<br>1.2. Confidence [45, 51]<br>1.2. Awareness [41, 45, 51]<br>1.3. Skills [41, 48]<br>1.4. Attitudes towards patients [41, 42, 45] | None | None |
| 2. Provider sense of community |  |  |  | **Organisation level**<br>2.1. Perceived support [41, 48]<br>2.2. Perception of being valued [41] |  |  |
| 3. Provider behaviour regarding trauma-informed care | None | None | **Organisation level**<br>3.1. Screening rates for traumatic experiences 25% →47% [50] | **Organisation level**<br>3.2. Uptake of self-care activities [41] | None | **Organisation level**<br>3.1. Screening for traumatic experiences [48]<br>3.2. Uptake of self-care activities [48] |
| 4. Patient readiness for disease management | None | None | **Individual-patient level**<br>4.2. Confidence in preventing and managing health conditions score | **Care-team level**<br>4.1. Feeling in control of treatment [42, 43, 48]<br>**Individual patient level**<br>4.2. Self-confidence [41-43] | None | None |
|  | Quantitative | Qualitative | Quantitative | Qualitative | Quantitative | Qualitative |
| | | | $\beta=0.213$, $p<0.001$ [49] | | | |
| 5. Patient satisfaction | None | None | **Organisation level**<br>5.1. Comfort and confidence in care score<br>$\beta=0.5284$, $p<0.001$ [49] | **Organisation level**<br>5.1. Comfort and confidence in care [41-44, 48]<br>5.2. Perceived support [41-44]<br>5.3. Satisfaction with services [41-44] | None | **Organisation level**<br>Satisfaction with services [48] |
| 6. Patient access to services | None | None | None | **Organisation level**<br>Access to services [41-44, 48] | None | None |
| 7. Provider and patient safety | None | None | None | **Organisation level**<br>7.1. Provider perceived safety [41]<br>7.2. Patient perceived safety [41-43] | None | None |
| **LONG-TERM HEALTH OUTCOMES, INDIVIDUAL-PATIENT LEVEL** |  |  |  |  |  |  |
| 8. Quality of life | None | None | Quality of life: 18 months $\beta=0.322$, $p<0.001$; 24 months $\beta=0.130$, $p<0.001$ [49] | None | None | None |
| 9. Chronic pain | None | None | Chronic pain: 18 months $\beta= - 0.194$, $p<0.001$; 24 months $\beta= - 0.078$, $p<0.001$ [49] | None | None | None |
| 10. Mental health | 10.1. Mental health symptoms score: 6-month effect size | None |  |  |  | None |
|  | Quantitative | Qualitative | Quantitative | Qualitative | Quantitative | Qualitative |
| | 95% CI crosses 0 [47]; 12-month 0.274 (95% CI -0.053 to 0.600) [53] | | 10.2. Depressive symptoms score: 18 months $\beta = -0.302$, $p < 0.001$; 24 months $\beta = -0.1223$, $p < 0.001$ [49]<br>10.3. PTSD score: 18 months $\beta = -0.305$, $p < 0.001$; 24 months $\beta = -0.123$, $p < 0.001$ [49] | 10.2. Depressive symptoms [42, 43] | 10.3. PTSD score: 6-month 95% CI crosses 0 [47]; 12-month 0.414 (95% CI 0.081 to 0.747) [53] | |
| 11. Substance use | 11.1. Alcohol use problem severity score: 6-month 95% CI crosses 0 [47]; 12-month 0.017 (95% CI 0.073 to 0.107) [53]<br>11.2. Drug use problem severity score: 6-month 95% CI crosses 0 [47]; 12-month 0.004 (95% CI -0.086 to 0.094) [53] | None | None | None | None | None |

Note. WCDVS, Women, Co-occurring Disorders and Violence Study. EQUIP, Equipping Primary Health Care for Equity study. ARISE, Aspire to Realize Improved Safety and Equity quality improvement programme.

The studies reported improvement in four out of seven psychological and behaviour outcomes and in two out of four health outcomes. Only three studies reported both psychological and health outcomes [41, 42, 48]. Although most interventions offered training and self-care activities for staff, no studies measured provider health and wellbeing. No studies reported adverse events/harm among patients or staff. No studies evaluated cost-effectiveness.

### Intermediate psychological and behavioural outcomes

We found limited evidence that some interventions may change organisational culture and create safe environments for patients and staff, potentially leading to improved perceived safety, patient disease management and access to services. These changes were measured through assessing organisational readiness to provide trauma-informed care, provider sense of community, patient readiness for disease management and access to services, and patient and provider perceived safety. However, the evidence for the effect on provider behaviour and patient satisfaction was conflicting.

#### Provider readiness

Four studies reported improvement in organisational readiness to provide trauma-informed care through promoting provider awareness, attitudes, confidence, and skills in the topic [41, 42, 45, 48]. Two qualitative evaluations reported that staff felt supported and valued [41, 48].

#### Provider behavior

In contrast, the evidence for change in collective behaviour and practices was conflicting. While the ARISE programme reported a 22% increase in screening rates for depression, substance abuse, and interpersonal violence [50], Dubay et al [48] found that in three US interventions, healthcare providers “had rich, nuanced views on the benefits and drawbacks of screening for trauma, which didn’t always neatly align with their organizations’ official policies.” Several interviewees called screening for trauma “controversial,” said they were “conflicted” or had “mixed” feelings about it or said “there are pros and cons (p.24)”. Similarly, while Bradley [41] found that staff used and appreciated well-being activities, Dubay et al [48] reported that “many interviewees said they did not adopt new self-care techniques after trainings, but appreciated grantees’ efforts to promote self-care (p.VII)”.

#### Patient psychological readiness for disease management

Two qualitative studies reported that patients felt in control of treatment [42, 48]. Four studies consistently reported improvement in patients’ confidence in managing health conditions and self-confidence [41, 42, 45, 48].

#### Patient satisfaction with services

Two studies found qualitative evidence for improved satisfaction with services [41, 42], while Dubay et al [48] reported that “some patients felt frustrated by high staff turnover and by the lack of staff diversity in some practices (p.33)”. **Patient access to services**. Three qualitative studies reported improved access to care through on-site provision or referrals to external organisations [41, 42, 48].

#### Provider and patient safety

Two qualitative studies reported improvement in perceived safety among patients and staff [41, 42] suggesting that the interventions created safe environments.

### Long-term patient health outcomes

We found limited and conflicting evidence with regard to change in some patient health outcomes. Three studies reported conflicting evidence for four health outcome domains with improvement in two, no effect in one, and mixed effects in one. The EQUIP study [45, 49] found strong evidence for improvement in patient quality of life, chronic pain, depression and PTSD symptoms at 18- and 24-month follow-ups. Brooks et al [42] found qualitative evidence for improvement in depressive symptoms. In contrast, the WCDVS reported no effect on mental health symptoms and severity of alcohol and drug problems; in this study PTSD symptoms remained unchanged at 6 months then improved at 12 months [47, 52, 53].

### Programme theories explaining intervention effects

We developed eight lines of argument for proposed integrated intervention mechanisms with limited evidence from five studies [41, 42, 45, 48, 52] (Table 2).

**Intervention worked as a package of components** in four studies [41, 42, 45, 48]. Two studies reported qualitative evidence explaining the challenging process of changing organisational readiness to provide trauma-informed care [45, 48, 51]. In the EQUIP study [45], this shift happened through “surfaced tensions that mirrored those in the wider community, including those related to racism, the impacts of violence and trauma, and substance use issues. Surfacing these tensions was disruptive but led to focused organizational strategies (p.1).” Similarly, Dubay et al [48] described tensions due to differing values and attitudes among staff groups. Further, the EQUIP study reported evidence linking engagement with the intervention and increased awareness and confidence among staff [45, 51]. Bradley [41] found qualitative evidence for the link between trauma-informed service and patients’ satisfaction and provider sense of community.

Brooks et al [42-44] reported some qualitative evidence suggesting that all components of the Trauma-informed Young Women’s Clinic contributed to patient satisfaction with services through improving access to healthcare for young women from deprived communities. Two studies reported some evidence for the link between intervention as a whole and outcomes *at the level of individual patient*. The WCDVS study reported that “intervention condition and programme elements” led to improvement in women’s trauma symptoms at 12 months (0.414 (95% CI 0.081 to 0.747) [53]. After using the Trauma-informed Women’s Clinic, some participants reported increased self-confidence, confidence in care, perceived safety and support, and improved mental health [42].

**Women-only space**, when services are provided by female providers to female patients, increased patient satisfaction with services through improving access to care, perceived safety and support, and self-confidence [41, 42]. However, Brooks et al [42] reported that “sometimes people get the wrong idea of the place, they think it’s for people that are man-haters (p.15)”.

**EQUIP care dose** was proposed as an indirect mechanism to patient health outcomes [49]. When patients perceived their care as more equity-oriented and trauma-informed, they felt more comfortable and confident in that care. This led to patients feeling more confident in their own ability to manage health problems. Over time, these psychological changes translated into better quality of life and less depression, trauma symptoms, and chronic pain.

**Staff education** was proposed as a mechanism for change in provider readiness. Two qualitative evaluations found that educating all staff about trauma-informed approach and self-care led to improvement in provider knowledge, skills [41, 48], and relationships [48].

**Tailoring staff education to organisational and wider context** was the EQUIP mechanism. Educational content and format were adapted to address clinic-specific needs, capacities, and priorities. Such tailoring contributed to staff feeling unified and to increased readiness for providing trauma-informed care [51].

**Staff self-care activities** contributed to staff perceived safety [41].

**Safe environment** created by the women-only policy, staff non-judgemental attitudes, and confidential services were reported as a link to patient safety and satisfaction through building trusting patient-provider relationships [41] and increasing patient perception of safety [42].

**Shared decision making** led to patient safety and satisfaction through education, feeling of control over treatment, and empowerment [42].

### Moderators

We developed lines of argument for the two overarching themes with seven sub-themes summarising factors that facilitated or hindered intervention effects: (i) contextual factors at the levels of wider political and economic environments, organisation (culture, resources), and individual patient (social determinants of health); (ii) intervention factors at the organisation level (intervention components, implementation process) (Figure 1, supplementary material Table S6).

#### Contextual moderators

Three qualitative studies identified political and economic conditions that could affect intervention effects [41, 45, 48]. These were relevant to values, regulatory, and financial regimes within the health system [41, 45, 48, 51]. Rigid policies, governance, and profit-driven business models made it difficult for healthcare providers to find time for training participation, self-care, and other organisational change activities [45, 48, 51]. Canadian and US providers acknowledged differing values between their organisations and the wider health system that acted as a barrier [45, 48]. Bradley [41] reported similar conflicting regulations and values in the UK third sector. EQUIP “participants noted the amplifying influence of other trauma-informed initiatives in the community” [51] (p.6).

Two studies described negative moderators at the organisation level: unsupportive culture with high pressure environment, disconnected leaders, hierarchical structure, differing values, and power imbalances [45, 48, 51]. In contrast, a supportive work environment and organisational values aligned with the principles of trauma-informed approach were described as positive moderators [51]. One study reported that their facility had limited capacity for changes in the physical environment [41].

One study described negative moderators at the individual patient level. Path analysis suggested that the EQUIP intervention was less effective for patients with experiences of intersecting structural violence (i.e., financial strain and discrimination) [49].

#### Intervention moderators

Barriers to the intervention implementation were reported most frequently. Four studies described poor engagement of some members of staff in intervention activities [48, 51], inadequate funding and dependence on project grants [41, 42, 51] as major barriers to sustainable organisational change. In contrast, two studies reported factors enabling successful workforce development leading to changes in the organisational culture. First, healthcare providers thought that collective learning through interprofessional conversations worked better than didactic methods [51]. Second, they highlighted the importance of the leadership buy-in [48]. Third, providers emphasised the importance of involving all staff in educational activities [48].

Two studies of services for women with a history of interpersonal violence found some evidence that components of the organisational change intervention can have a modifying effect. The hierarchical linear modelling in the WCDVS produced conflicting results. While receiving integrated counselling for trauma, mental health, and substance abuse resulted in better patient health outcomes, receiving more study services resulted in less improvement [53]. Bradley [41] quoted “one member of staff emphasised the importance of prioritising staff well-being as equal to the support provided to women (p.16)”.

## Discussion

### Principle findings

This mixed methods systematic review of six non-randomised studies which assessed eight models of trauma-informed organisational change interventions in primary care and community mental healthcare found limited and conflicting evidence for their effects on patient and healthcare provider psychological, behavioural, and health outcomes. Healthcare organisations tailored different models of trauma-informed organisational change to their needs, abilities, and preferences. The most common components included an allocated budget, ongoing training and support for all staff, identification and treatment for trauma, and evaluation. Four studies reported improvement in provider readiness to deliver trauma-informed care, and improvement in their sense of community. However, two studies reported that only some providers used self-care activities and screened for traumatic experiences.

Four studies reported some improvement in patient readiness for disease management and access to services; however, the evidence for patient satisfaction was conflicting. Two studies found that patients and providers felt safe. While one study reported some improvement in patient quality of life and chronic pain, three studies reported conflicting findings regarding effect on mental health symptoms, and one found no effect on alcohol and drug problem severity. No studies measured adverse events/harm, cost effectiveness, or staff health and well-being. The limited evidence for programme’ mechanisms suggested that interventions may work either as a whole or through separate components – staff education tailored to the local context, self-care activities for staff, safe environments, and shared treatment decision making. We identified contextual and intervention factors that may moderate intervention effects. Contextual moderators included health system values, policies, governance, and business models, wider trauma-informed programmes, organisational culture, and patient social determinants of health. Intervention moderators required buy-in and engagement from all staff, collective learning through interprofessional conversations, equal attention to well-being of staff and patients, and sustainable funding.

Our first important finding is that the empirical evidence base for the effectiveness of trauma-informed organisational change interventions in primary care and community mental healthcare is very limited. Despite exhaustive searches, we only identified three non-randomised quantitative studies and three qualitative service evaluations of different intervention models. One of the reasons for the evidence gap could be the methodological challenges of evaluating organisational change interventions within complex health systems. However, the literature offers varied tools and guidance on how to monitor and evaluate trauma-informed organisational change interventions [32, 35, 56, 57]. It is possible that some evaluation reports have not been made public.

Our second important finding is that despite heterogeneity in the included models and evaluation designs, we found comparable domains for intervention components, outcomes, mechanisms, and moderators. By mapping intervention components on the SAMHSA trauma-informed approach framework [9], we showed that all eight intervention models were built on the *4Rs* assumptions and all included components within the same domains: (i) budget allocation, (ii) training and workforce development, (iii) identification and/or response to violence and trauma, and (iv) evaluation of the organisational change. Such similarities can explain convergence of effects when these were detected. They may work through the same mechanisms of changing provider readiness, sense of community, and safety to changes in patient readiness, satisfaction, safety, and health. Our findings on outcome domains and possible mechanisms are in line with recent systematic reviews of trauma-informed interventions at the organisational level that did not include any of our studies [7, 22]. Both reviews found conflicting effects on provider readiness and practices regarding provision of trauma-informed care, and service user perception of care. None of our studies used validated measures for evaluating organisational readiness, culture, and performance identified by Melz et al [7]. The overlapping mechanism for increased provider readiness was staff education and ongoing support.

Our third important finding about increased perceived safety and sense of community among healthcare providers indicates positive changes in organisational environments, relationships, and culture which may facilitate and support subsequent changes in clinical practices. This finding supports the recommendation for the organisation domain of the trauma-informed approach to be the pre-condition which enables and helps sustain trauma-informed changes in clinical practices by individual healthcare providers [8, 9].

Our fourth finding supports the proposition that a universal trauma-informed approach does not have to include screening component to improve patients’ experiences and outcomes. In our review, the five models which included screening for trauma (WCDVS, ARISE programme, Women’s HIV Clinic, Montefiore Medical Group, One-stop-shop Women’s Clinic) and the three models that did not include screening for trauma (EQUIP, Trauma-informed Young Women’s Clinic, and Sanctuary Model) reported improvement in some patient outcomes (readiness for disease management, safety, and health). No studies explored the mechanisms linking screening to health outcomes or harm among patients and staff.

Providers had conflicting views on the acceptability and feasibility of screening all patients for traumatic experiences. This finding on the uncertain evidence for the effectiveness and safety of screening for traumatic experiences is in line with the recent systematic reviews [7, 22].

### Strengths and limitations

We conducted a methodologically robust systematic review with two reviewers working in parallel at each stage. Our rigorous search strategy included both peer-reviewed and grey literature without language restrictions other than inclusion of an English abstract. This resulted in a global view of trauma-informed organisational change interventions in primary and community mental healthcare. Additionally, we contacted and received responses from study authors to identify other relevant studies and to clarify information regarding data extraction and quality appraisal. A methodological limitation is that we used search terms based on trauma-informed terminology introduced in early 2000, which meant that earlier studies meeting the inclusion criteria may have been excluded, as they were not labelled as ‘trauma-informed’. We addressed this limitation through seeking input from our public and professional advisory groups when designing the search strategy. We involved people with lived experience and professionals in different stages of the review to ensure that our findings are relevant and beneficial to them. By using a logic model to map review findings, we produced findings that are understandable to healthcare providers and policy makers. Exclusion of papers without an English abstract might have resulted in missing relevant studies reported in other languages.

The evidence we found is very limited and uncertain due to the small number and non-randomised designs of the primary studies. That said, the included studies were generally characterised by good sample size. Non-randomised studies provide weaker evidence of causal effects of interventions on outcomes. Although we tried to hypothesise causal links through mapping onto our logic model, these are assumptions supported by six studies at the bottom of the hierarchy of evidence and further high-quality research in this area is warranted.

### Implications for policy, practice, research

Any generic framework for trauma-informed approach should be contextually tailored for organisational needs, abilities, and preferences. If primary care and community mental healthcare organisations integrate trauma-informed assumptions and principles across at least six implementation domains, they may change organisational culture and create safe environments for staff and patients potentially leading to improvement in patient disease management and satisfaction, access to services, quality of life and chronic pain. Every trauma-informed organisational change intervention should have funding for an embedded evaluation and agreement on target outcomes and measures that are evidence based and theoretically informed. Future research exploring trauma-informed approaches in primary care and community mental healthcare should include randomised designs and validated measures to capture changes across individual, team, organisation, wider system levels and enable meta-analysis. Studies should also evaluate adverse events/harm, provider health, and cost-effectiveness.

## Conclusions

Trauma-informed organisational change interventions in primary care and community mental healthcare may improve provider readiness and sense of community, patient readiness for disease management and access to services, provider and patient safety, and some patient health outcomes, but the evidence is very limited and conflicting.

## Supporting information

Supplementary material all files

## Data Availability

The quantitative and qualitative data supporting this systematic review were extracted from previously reported studies, which have been cited. The processed data are included within the article and supplementary material file.

## Data availability

The quantitative and qualitative data supporting this systematic review are from previously reported studies, which have been cited. The processed data are included within the article and supplementary material files.

## Acknowledgments

We are grateful to the members of the public and professional advisory groups who represented people with lived experience of violence and trauma and professionals who plan, fund, and deliver health services. Their advice and critical feedback on study design, findings, and outputs were invaluable.

## Supplementary material

Table S1. Trauma-informed approach at the organisational (synonym system) level: definitions and core components

Table S2. Example search strategy

Table S3. Methodological quality of included studies

Table S4 Methodological quality for the overall study design, data collection and analysis

Table S5. Factors affecting effectiveness of trauma-informed organisational change interventions

