## Supplementary material all files for "Trauma-informed approaches in primary healthcare and community mental healthcare: a mixed methods systematic review of organisational change interventions"

Table S1. Trauma-informed approach at the organisation level: definitions and core components

| **Year**  **Authors**  **Document** | **Definition of trauma** | **Definition of trauma-informed approach** | **Assumptions** | **Philosophy and principles** | **Implementation domains** |
| --- | --- | --- | --- | --- | --- |
| 2001. Harris and Fallot [8]  Using trauma theory to design service systems | Current or past sexual or physical abuse trauma. Trauma is not a single discrete event but rather an organising experience that forms the core of the individual’s identity. | Trauma-informed services are not designed to treat symptoms or syndromes related to sexual or physical abuse. Regardless of their primary mission their commitment is to provide services in a manner that is welcoming and appropriate to the special needs of trauma survivors. | 1. To know the history of past and current abuse  2. To understand the impact of violence and victimization  3. Accommodate the vulnerabilities of trauma survivors  4. Facilitate consumer participation in treatment  5. Avoid retraumatising and revictimizing consumers | Paradigm shift within service delivery systems:  1. Understanding the impact of trauma  2. Holistic approach  3. Strength-based approach to prevent problematic behaviour  4. Collaborative clinical decision making  5. Open and genuine collaboration between providers  6. Trust and safety | Certain conditions need to be in place for a trauma-informed system to be established. Those conditions reflect the structure and culture of the organisation, and they predate any actual changes in clinical services:  1. Administrative commitment to change  2. Training and education for all staff  3. Sensitive hiring practices  4. Review of policies and procedures  5. Universal screening, trauma assessment |
| 2014. Substance Abuse and Mental Health Services Administration (SAMHSA), US [9]  SAMHSA’s concept of trauma and guidance for a trauma-informed approach | Individual trauma results from an **event**, series of events, or set of circumstances that is **experienced** by an individual as physically or emotionally harmful or life threatening and that has lasting adverse **effects** on the individual’s functioning and mental, physical, social, emotional, or spiritual well-being. | A strength-based service delivery approach that is grounded in an understanding of and responsiveness to the impact of trauma, that emphasise physical, psychological, and emotional safety for both providers and survivors, and that creates opportunities for survivors to rebuild a sense of control and empowerment. | The *four Rs*:  1. Realisation of the impact of trauma and understanding paths to recovery  2. Recognition of signs and symptoms of trauma  3. Response by integrating knowledge about trauma into policies, procedures, and practices  4. Resist re-traumatisation. | 1. Safety  2. Trustworthiness and transparency  3. Peer support  4. Collaboration and mutuality  5. Empowerment, voice, and choice  6. Cultural, historical, and gender sensitivity | 1. Governance and leadership  2. Policy  3. Physical environments  4. Engagement and involvement  5. Cross sector collaboration  6. Screening, assessment, treatment services  7. Training and workforce development  8. Progress monitoring and quality assurance  9. Financing  10. Evaluation |
| 2017. NHS Education for Scotland and Scottish Government, UK [12]  NHS Education for Scotland. Transforming psychological trauma: a knowledge and skills framework for the Scottish workforce | SAMHSA definition | The Transforming Psychological Trauma framework | 1. Recognises prevalence and impact of trauma  2. Respond  3. Prevent re-traumatisation | 1. Safety  2. Choice  3. Collaboration  4. Trust  5. Capacity-strengthening approach  6. Acknowledges rights  7. Access to treatment | 1. Training for all workforce  2. Staff well-being |
| 2017. Centre for Health Care Strategies, Inc., US [10]  What is trauma-informed care? | SAMHSA definition. Examples of trauma include physical, sexual, and emotional abuse, childhood neglect, having a family member with mental health or substance abuse disorder, violence in the community and poverty and systemic discrimination. | SAMHSA definition. | The SAMHSA *four Rs* | 1. Patient empowerment  2. Choice  3. Collaboration  4. Safety  5. Trustworthiness | Organisational reform precedes the adoption of trauma-informed clinical practices.  1. Leading and communication about the transformation  2. Engaging patients in organisational planning  3. Training clinical and non-clinical staff  4. Creating safe environment  5. Preventing secondary traumatic stress in staff  6. Hiring a trauma-informed workforce  Clinical practices:  7. Involving patients in the treatment process  8. Screening for trauma  9. Training staff in trauma-specific treatment approaches  10. Engaging referral services and partnering organisations |
| 2018. The Women’s Mental Health Taskforce, UK [5]  The Women's Mental Health Taskforce. Final report | Violence, abuse, poverty, and inequality. | Gender- and trauma-informed approach  refers to an approach by which organisations operate with an awareness of trauma and its impact and avoid re-traumatisation | 1. Recognise the impact of trauma  2. Avoid re-traumatisation for staff or service users  3. Identify recovery from trauma as a primary goal | 1. Whole organisation approach  2. Equal access  3. Recognition and response to violence and trauma  4. Respectful, compassionate, trustful relationships  5. Safety  6. Engagement of service users  7. Capacity- strengthening, empowering approach  8. Holistic approach  9. Gender-informed approach. | Not reported |
| 2018. Public Health Agency of Canada [13]  Trauma and violence-informed approaches to policy and practice | Trauma-informed approaches are familiar to many organizations and service providers. Recently, this term has been expanded to include "and violence", an important change in the language which underscores the connections between trauma and violence | Trauma and violence-informed approaches are policies and practices that recognize the connections between violence, trauma, negative health outcomes and behaviours. These approaches increase safety, control and resilience for people who are seeking services in relation to experiences of violence and/or have a history of experiencing violence | Recognise connections between violence, trauma, behaviour, health | 1. Understand trauma and violence, and their impacts on peoples' lives and behaviours  2. Create emotionally and physically safe environments  3. Foster opportunities for choice, collaboration, and connection  4. Provide a strengths-based and capacity-building approach to support client coping and resilience | 1. Policies and practices  2. Environments |
| 2019. Gerber [11]  Trauma-informed healthcare approaches: a guide for primary care | SAMHSA definition of individual trauma, complex trauma, historical trauma, structural violence. | SAMHSA definition  Trauma-informed systems of care strive to become vicarious trauma-informed by attending pro-actively and compassionately to the vicarious trauma of healthcare providers and staff. | Not reported | SAMHSA principles  AND  *Four Cs* of clinical care:  1. Calm  2. Contain  3. Care  4. Cope (p. 33) | Not reported |
| 2021. NHS England and NHS Improvement, UK [14]  A good practice guide to support implementation of trauma-informed care in the perinatal period | SAMHSA definition | SAMHSA definition | Not reported | 1. Compassion and recognition  2. Communication and collaboration  3. Consistency and continuity  4. Recognising diversity and facilitating recovery | 1. Leading and communicating change  2. Staff training  3. Supervision and peer support  4. Co-production and service co-design  Evaluation and culture of improvement |

Note. SAMHSA, the Substance Abuse and Mental Health Services Administration is a branch of the U.S. Department of Health and Human Services. US, United States. UK, United Kingdom. NHS, National Health Service.

Table S2. Example search strategy

Database: Ovid MEDLINE(R) 1946 to present

| **#** | **Searches** | **Results** |
| --- | --- | --- |
| 1 | (trauma inform* or trauma-inform* or trauma focus* or trauma-focus* or trauma responsive* or trauma sensitive or trauma-sensitive or trauma services or complex trauma or trauma-base* or trauma base*).ab,ti. | 3556 |
| 2 | (trauma-inform* and (approach* or care or practi? or intervention* or system*)).ab,ti. | 1185 |
| 3 | (trauma-inform* and (measure or survey or questionnaire or checklist or assessment or evaluation)).ab,ti. | 445 |
| 4 | or/1-3 | 3556 |
| 5 | (adverse childhood event$ or adverse childhood experience$ or psychological* informed environment$).ab,ti. | 2404 |
| 6 | 4 or 5 | 5800 |
| 7 | (child or child* or children).mp. | 2538736 |
| 8 | child/ or children/ or infant/ or adolescent/ | 3288214 |
| 9 | or/7-8 | 3801413 |
| 10 | 6 not 9 | 1932 |

Table S3. Models of trauma-informed organisational change interventions in primary care and community mental healthcare

| **SAMHSA Implementation domains** [9] | **Women, Co-occurring Disorders, and Violence Study,** McHugo 2005  [47, 52, 53] | **Equipping Primary Health Care for Equity (EQUIP) model,** Browne 2018  [45, 46, 49, 51] | **Aspire to Realize Improved Safety and Equity (ARISE) quality improvement programme,** Kimberg 2019  [50] | **Advancing Trauma Informed Care initiative,**  Dubay 2018 [48] | | | | | **Trauma-informed Young**  **Women’s Clinic,** Brooks 2017  [42-44] | **One-stop-shop Women’s Centre (Trauma-informed service system model),** Bradley 2020  [41] | **Interventions models (n=8)** |
| --- | --- | --- | --- | --- | --- | --- | --- | --- | --- | --- | --- |
|  |  |  |  | **Women’s HIV Clinic (Trauma-informed Primary Care model)** | | **Montefiore Medical Group** | **Family Health Services (Sanctuary model)** | |  |  |  |
| **1. Governance and leadership** | Not reported | Engagement of clinical/administrative leaders/managers in consultations with clinicians and other staff to conduct local needs assessment, select 3-5 priorities, and develop plan for addressing each priority. | The quality improvement team includes the Director of Primary Care, Behavioural Health, the Primary Care Director of Population Health and Quality, the quality improvement coordinator, and senior clinicians. | Not reported | | Not reported | Complies with the Sanctuary Model certification. Steering Committee, monthly meetings open to all staff. | | Organisation mission statement reflects a social model of health care and includes commitment to providing trauma-informed care. | Organisation mission statement includes commitment to providing trauma-informed care. | **5** |
| **2. Written policies and procedures** | Not reported | New harm reduction policies and practices for supporting patients with substance use issues. | System-wide quality improvement priorities, aligned with state and national performance metrics and associated incentives for depression and alcohol/substance use.  Standardise, team-based workflow.  Screening and response protocols. | Not reported | | Not reported | Not reported | | Written policy recognises impact of gender-based violence, promotes women’s role in service delivery, commitment to staff development | Not reported | **3** |
| **3. Physical environment** | Women only space. | Removed sign ‘Check in or you will lose your appointment’ from the reception desk.  Opened clinic doors 30 min earlier so that patients could wait indoors to book appointments.  Repurposed a section of the waiting room to create a child-friendly space for patients with small children. | Not reported | Redesigned waiting room to be more calming for staff and patients.  Chair sessions with massage therapist for patients.  Service dog for patients to pet.  Free food for patients.  colouring pages.  Calming music. | | Waiting room posters about the importance of behavioural health treatment for traumatic experiences | Natural light in many rooms.  Open space. | | Women only space.  Located in an older house in the community.  Comfortable furniture.  Service users’ art and posters affirming women’s rights.  Consultation length is tailored to patient needs.  Childcare during clinic activities. | Women only space.  Female only staff.  Variety of services in one location.  Fully staffed creche for childcare during activities.  Non-clinical reception and waiting area.  Open safe space.  Empowering and motivational quotes, bright paintings.  Light and comfortable room for group work.  Physical environment is regularly assessed through ‘walk-throughs’ and reflective practices. | **7** |
| **4. Engagement and involvement of people with lived experience** | Service users and  HCP with lived experience helped design and/or deliver trauma-specific interventions. Sourcebook for developing partnerships between HCPs and people with lived experience. | Hired and integrated an Indigenous Elder to participate in clinic activities. | Patient advisory board.  Educational message on the screening tool to precede the questions and a checklist of coping behaviours and resilience factors to facilitate patient-centred conversations about preferred treatment. | Stakeholder group including four patients and representatives from each department meets monthly. The group provides feedback, helps design and implement organisational change. | | Not reported | Not reported | | Person-centred care.  Shared decision making. | Lived experience panels and forums. Recruit staff with lived experience.  Varied activities, food sharing, creative projects. | **6** |
| **5. Cross sector collaboration** | Linkages across agencies, all services were comprehensive, integrated, and trauma informed. | Developed new working relationship with the local Indigenous community. | Formalised partnerships with a community-based domestic violence agency, a legal aid organisation, a trauma-specific treatment organisation, and a national non-profit violence prevention resource centre. | Partnerships. | | Not reported | Not reported | | Referrals from the local high school and other services supporting young people experiencing adversity.  Partnership with community youth-oriented services. | Engaged with local partnerships and companies to improve service and supplies for women.  Collaborative partners are trauma-informed. | **6** |
| **6. Screening, assessment, treatment services** | Screening, assessment. Simultaneous and coordinated provision of substance abuse, mental health, and trauma services.  On-site trauma-specific group intervention TREM (Trauma Recovery and Empowerment Model).  Peer groups, advocates, facilitators, drop-in clinic. | Integrated trauma- and violence-informed approaches into the routine provision of care.  Chronic pain group. | Screening all patients concurrently for depression, alcohol/substance use and interpersonal violence.  Single screening tool and pathway for depression, alcohol/substance use, and interpersonal violence.  On-site cognitive behavioural interventions and interpersonal violence advocate.  Referrals to mental health and substance abuse services in the community. | Screening for interpersonal violence, PTSD, substance abuse, chronic pain.  On- and off-site psychosocial services.  Collocate a licensed clinical social worker to offer on-site therapy.  Group intervention on skill building for patients who aren’t ready for traditional talk therapy. | | Screening for adverse childhood experiences.  On-site behavioural health counsellors. | On-site talking therapy, creative art therapies.  On-site social workers, behaviour health counsellors, nutritionist, fitness centre, yoga and mindfulness classes for patients. | | Drop-in appointments with a nurse, counsellor, and general practitioner.  Trauma-sensitive gynaecological care.  Drop-in facilitated art group with  peer facilitators. | Universal screening for histories of trauma.  On-site drop-in support, one to one support, well-being groups, trauma-specific interventions, tailored rehabilitation support. | **8** |
| **7. Training and workforce development** | Community support specialists trained in trauma, mental health, and substance abuse. | All staff training: 2-and 8-hour workshops, online 8-hour self-directed programme.  Strategy and support for vicarious trauma among staff.  Trauma champion practice consultant. | Staff training | All staff training.  Three half-day initial mandatory trainings, three 1.5-hour training for new hires.  Weekly staff meetings and pre-clinic meetings.  Outside trainer attends once a month to discuss trauma with staff.  Trauma champion clinical social worker. | | All staff training with mandatory initial training.  New Critical Incident Management Team to counsel practices after traumatic event (e.g., a shooting).  Quarterly learning collaboratives for teams.  Role-specific, in-person trainings led by therapists.  Relaxation hotline for staff. | All-staff training with mandatory initial training.  All-staff meetings.  Mindfulness and yoga classes for staff.  2 hours a month for staff self-care.  An “Undoing Racism” committee.  Reflective supervision. | | Staff supervision sessions with a trauma specialist counsellor.  Self-care for staff. | Trauma champion in each team.  Hiring practices.  All staff training including self-care and well-being, specialised training for staff wishing to deliver trauma-responsive and trauma-specific services.  Staff wellbeing days.  Trauma-informed supervision, clinical supervision. | **8** |
| **8. Progress monitoring and quality assurance** | Not reported | Trauma champion assisted staff to evaluate the progress and adjust the plan. | The Behavioral Health Vital Signs (BHVS) performance metric, BHVS screening tool and quality improvement process.  Data visualisation dashboards for tracking progress and a monthly meeting for clinics to share best practices.  Electronic health record templates and data. | Stakeholder group provides feedback.  Ongoing monitoring and evaluation. | | Tracking progress by interviewing or surveying staff. | Tracking progress by interviewing or surveying staff. | | Solicit feedback from service users.  Regular staff check in with management.  Annual planning days that encourage staff feedback. | Monthly meetings of the organisation wide reflective group focussed on the question – How trauma informed are we? | **7** |
| **9. Financing** | Grant from SAMHSA. | Grant from the Canadian Institute of Health Research, each clinic received $10,000 for 24 months. | Federal funds for on-site behavioural health clinicians.  Incentivised performance metrics for depression and alcohol and substance abuse disorders.  ARISE grant. | 24-month grant from the Robert Wood Johnson Foundation. | | | | | Medicare bulk billing and partnership with community youth-oriented services.  Free for patients. | Grants from the National Lottery, fundraising. | **8** |
| **10. Evaluation** | 12-month quantitative study | 24-month mixed methods study | Quantitative evaluation of the quality improvement programme | Cross-sectional qualitative service evaluation | | | | | Cross-sectional qualitative service evaluation | Cross-sectional qualitative service evaluation.  Lived experience forums, feedback opportunities in group work, comments box in the centre. | **8** |
| **Domains, n** | **7** | **10** | **9** | **8** | **6** | | | **7** | **10** | **9** |  |

Note. Studies listed in chronological order. SAMHSA, the Substance Abuse and Mental Health Services Administration. ACEs, adverse childhood experiences.

Table S4. Methodological quality of included studies

| **Study ID, reports** | **Screen** | | **Qualitative** | | | | | **Quantitative** | | | | | **Mixed methods** | | | | | **Total** |
| --- | --- | --- | --- | --- | --- | --- | --- | --- | --- | --- | --- | --- | --- | --- | --- | --- | --- | --- |
|  | **S1** | **S2** | **1.1** | **1.2** | **1.3** | **1.4** | **1.5** | **3.1** | **3.2** | **3.3** | **3.4** | **3.5** | **5.1** | **5.2** | **5.3** | **5.4** | **5.5** | **%Yes** |
| Browne 2018 [45, 49, 51] | Yes | Yes | Yes | Yes | Yes | Yes | Yes | Yes | Yes | Yes | Yes | Yes | Yes | Yes | Yes | No | Cannot tell | 88 |
| Kimberg 2019 [50] | No | Cannot tell | NA | NA | NA | NA | NA | Yes | Yes | No | No | Cannot tell | NA | NA | NA | NA | NA | 29 |
| Dubay 2018 [48] | Yes | Yes | Yes | Yes | Yes | Yes | Yes | NA | NA | NA | NA | NA | NA | NA | NA | NA | NA | 100 |
| Brooks 2018 [42-44] | Yes | Yes | Yes | Yes | Yes | Yes | Yes | NA | NA | NA | NA | NA | NA | NA | NA | NA | NA | 100 |
| Bradley 2020 [41] | Yes | Yes | Yes | Yes | Yes | Yes | Yes | NA | NA | NA | NA | NA | NA | NA | NA | NA | NA | 100 |
| McHugo 2005 [47, 52, 53] | Yes | Yes | NA | NA | NA | NA | NA | Yes | Yes | Yes | Cannot tell | Cannot tell | NA | NA | NA | NA | NA | 71 |

Table S5. Methodological quality for the overall study design, data collection and analysis

|  | **Yes** | | **No** | | **Cannot tell** | | **Total assessed** | |
| --- | --- | --- | --- | --- | --- | --- | --- | --- |
|  | **N** | **%** | **N** | **%** | **N** | **%** | **N** | **%** |
| **Screening questions**  S1. Are there clear research questions? | 5 | 83 | 1 | 17 | 0 | 0 | 6 | 100 |
| S2. Do the collected data allow to address the research questions? | 5 | 83 | 0 | 0 | 1 | 17 | 6 | 100 |
| **Qualitative study component**   1.1. Is the qualitative approach appropriate to answer the research question? | 4 | 100 | 0 | 0 | 0 | 0 | 4 | 100 |
| 1.2. Are the qualitative data collection methods adequate to address the research question? | 4 | 100 | 0 | 0 | 0 | 0 | 4 | 100 |
| 1.3. Are the findings adequately derived from the data? | 4 | 100 | 0 | 0 | 0 | 0 | 4 | 100 |
| 1.4. Is the interpretation of results sufficiently substantiated by data? | 4 | 100 | 0 | 0 | 0 | 0 | 4 | 100 |
| 1.5. Is there coherence between qualitative data sources, collection, analysis and interpretation? | 4 | 100 | 0 | 0 | 0 | 0 | 4 | 100 |
| **Quantitative non-randomised study component**  3.1. Are the participants representative of the target population? | 3 | 100 | 0 | 0 | 0 | 0 | 3 | 100 |
| 3.2. Are measurements appropriate regarding both the outcome and intervention (or exposure)? | 3 | 100 | 0 | 0 | 0 | 0 | 3 | 100 |
| 3.3. Are there complete outcome data? | 2 | 67 | 1 | 33 | 0 | 0 | 3 | 100 |
| 3.4. Are the confounders accounted for in the design and analysis? | 1 | 33 | 1 | 33 | 1 | 34 | 3 | 100 |
| 3.5 During the study period, is the intervention administered (or exposure occurred) as intended? | 1 | 33 | 0 | 0 | 2 | 67 | 3 | 100 |
| **Mixed methods study component**  5.1. Is there an adequate rationale for using a mixed methods design to address the research question? | 1 | 100 | 0 | 0 | 0 | 0 | 0 | 100 |
| 5.2. Are the different components of the study effectively integrated to answer the research question? | 1 | 100 | 0 | 0 | 0 | 0 | 0 | 100 |
| 5.3. Are the outputs of the integration of qualitative and quantitative components adequately interpreted? | 1 | 100 | 0 | 0 | 0 | 0 | 0 | 100 |
| 5.4. Are divergences and inconsistencies between quantitative and qualitative results adequately addressed? | 0 | 0 | 1 | 100 | 0 | 0 | 0 | 100 |
| 5.5. Do the different components of the study adhere to the quality criteria of each tradition of the methods involved? | 0 | 0 | 0 | 0 | 1 | 0 | 0 | 100 |

Note. Methodological quality was appraised with the Mixed Methods Appraisal Tool (MMAT) version 2018. NA not applicable.

Table S6. Factors affecting effectiveness of trauma-informed organisational change interventions

| **Factor theme** | **Definition** | **Discussed by (study ID, reports)** | **Supporting quote** |
| --- | --- | --- | --- |
| 1. Contextual factors | Hindering or enabling conditions under which trauma-informed organisational change intervention is enacted. | Bradley 2020, Browne 2018, Dubay 2018  [41, 45, 48, 49, 51] |  |
| 1.1. Political and economic environments | Governance, financial, payment regulations and health system values. | Bradley 2020, Browne 2018, Dubay 2018 [41, 45, 48] | 1. “One interviewee said: “Trauma-informed systems is focused on relationships and building or repairing relationships, which takes time and processes. That is part of the work that needs to happen. But at the same time, we all have goals and need to see a certain number of clients, and we all need to process a certain number of contracts, whatever our own department is responsible for.” Clinical staff in three organizations said it was difficult to participate in trauma-informed efforts when most of their day was spent seeing patients and the business model of the organization depends on revenues from treating patients.” [48]  2. “Staff members realized that their organizational mandates to pay attention to equity were somewhat ‘out of step’ with values driving the larger health care system, and that the positioning of the clinics within the wider health care system limited their possibilities to provide EOHC [equity-oriented health care]” [45] |
| 1.2. Wider trauma-informed movement | Parallel trauma-informed initiatives in the community, health system. | Browne 2018 [51] | 3. “Participants noted the amplifying influence of other trauma-informed initiatives in the community, with one saying: “There’s been lots of information and training within the community, the community organizations just like us and I think that it’s making a difference in how we work. (601, Counselor)” [51] |
| 1.3. Organisational culture | Organisational values, psychological environment, leadership style, receptiveness to principles of trauma-informed care. | Browne 2018, Dubay 2018 [45, 48, 51] | 4. “I’m not convinced that administration has any concept of how swamped we’re all feeling. This lack of communication from the board, and this is what I’m hearing, and I’m kind of seeing it, this lack of involvement from the board has caused … low morale. Sometimes I feel like there’s a bit of a disconnect between the primary care providers, like what happens down here, like providing care for the patients, and then what goes on upstairs [in the administration area]?” [45]  5. “One primary care physician observed cultural differences between her professional field and the behavioral health care field, which she believed explained why some primary care staff were less interested in some self-care activities. This interviewee felt that primary care staff did not have the luxury of thinking about things like yoga because they already had enough trouble making time to eat lunch, with their hectic work schedules. A behavioral health provider at another organization agreed: “Medical doctors are funny people. There’s not a lot of self-care. In the mental health world, we talk about self-care all the time. In the medical world, it seems far less talked about.” [48]  6. “organizational hierarchy was a barrier to becoming trauma-informed. Hierarchy challenges varied across respondents and organizations and included lack of racial diversity in leadership roles and hiring practices; power dynamics among physicians, therapists, nurses, and medical assistants; inconsistent supervisory support to give voice and innovate around trauma-informed efforts; and insufficient incorporation of patient voices, needs, and requests into the practice.” [48]  7. “Multiple participants described how their particular clinic context helped or hindered their efforts at TVIC [trauma- and violence-informed care]. Clinic mandates and culture often aligned with TVIC concepts. However, the extent to and ways in which TVIC was taken up in each clinic was influenced by the existing interprofessional culture, tensions and power dynamics. One leader described how the culture in her team supported TVIC practice: We have a work environment that allows us to be supportive of each other so that we can have the emotional reserves to be able to provide trauma informed care. I think we’re flexible: if you have somebody that comes in that’s in a particular trauma or crisis, our colleagues are always really good about accommodating that time. (502, Administrator)” [51] |
| 1.4. Organisational resources | Facility characteristics and material conditions for trauma-informed organisational changes. | Bradley 2020 [41] | 8. “The Women’s Centre is based in a listed building and as such, it is acknowledged that renovations to the physical space may not be straight forward.” [41]. |
| 1.5 Patient characteristics | Individual patient characteristics that hindered or facilitated intervention effects. | Browne 2018 [49] | 9. Patient “financial strain and experiences of discrimination had significant negative effects on all health outcomes.” [49] |
| 2. Intervention factors | Hindering or enabling factors in the intervention design and implementation process. | Bradley 2020, Brooks 2017, Dubay 2018, Browne 2018, McHugo 2005 [41, 42, 48, 51, 53] |  |
| 2.1. Implementation process | Barriers and enablers to intervention application: staff engagement in intervention activities, funding for organisational change. | Bradley 2020, Brooks 2017, Dubay 2018, Browne 2018 [41, 42, 48, 51] | 10. “Another barrier was resistance or pushback from subsets of staff (which varied by organization but often spanned many staff levels), both during and after trainings, to changes such as increasing the use of an ACEs [Adverse Childhood Experiences] screening questionnaire and participating in staff self-care strategies such as meditation and yoga classes.” [48]  11. “most organizations relied on funding from this and other grants to implement and enhance their trauma-informed efforts; these opportunities may not be available to all organizations or providers.” [48] |
| 2.2. Staff education | Barriers and enablers for staff education: interactive learning, buy-in from leadership, all staff, accessibility | Dubay 2020, Browne 2018 [48, 51] | 12. “A key finding in this study was that staff at both clinics found the interprofessional conversations more impactful than the didactic education. These conversations provided staff with the opportunity to learn together, to learn about one another’s perspectives on violence and trauma in their practices, and to take a level of collaborative action together that was unprecedented.” [51]  13. “Staff at nearly all organizations felt that leadership buy-in was one of the most important elements and that when leadership did not believe in the value of a training, neither would staff from that organization. And for trainings to happen, leadership needed to believe that they were worth the money and time away from patient care. At some organizations, only a limited number of staff were excused from clinical duties to attend some trainings, which obviously limited the reach of these trainings.” [48] |
| 2.1. Intervention components | Intervention components that modified intervention effect: integrated services, varied services, equal attention to supporting patients and staff. | Bradley 2020, McHugo 2005 [41, 53] | 14. “Analysis of key program elements demonstrated that integrating substance abuse, mental health, and trauma-related issues into counseling yielded greater improvement, whereas the delivery of numerous core services yielded less improvement relative to the comparison group.” [53]  17. “One member of staff emphasised the importance of prioritising staff well-being as equal to the support provided to women.” [41] |
